# Predicting Eating Disorder and Anxiety Symptoms Using Anorexia Nervosa and Obsessive-Compulsive Disorder Polygenic Scores

**DOI:** 10.1101/2020.07.02.20142844

**Authors:** Zeynep Yilmaz, Katherine Schaumberg, Matt Halvorsen, Erica L. Goodman, Leigh C. Brosof, James J. Crowley, Anorexia Nervosa Genetics Initiative, Eating Disorders Working Group of the Psychiatric Genomics Consortium, Tourette Syndrome/Obsessive-Compulsive Disorder Working Group of the Psychiatric Genomics Consortium, Carol A. Mathews, Manuel Mattheisen, Gerome Breen, Cynthia M. Bulik, Nadia Micali, Stephanie C. Zerwas

**Author notes:** Corresponding author: Nadia Micali, MD. Department of Psychiatry, Faculty of Medicine, University of Geneva, HUG, Geneva, Switzerland. Co-senior authors (Nadia Micali, Stephanie C. Zerwas).

## Abstract

Clinical, epidemiological, and genetic findings support an overlap between eating disorders, obsessive-compulsive disorder (OCD), and anxiety symptoms. However, little research has examined the role of genetic factors in the expression of eating disorders and OCD/anxiety phenotypes. We examined whether the anorexia nervosa (AN), OCD, or AN/OCD transdiagnostic polygenic scores (PGS) predict eating disorders, OCD, and anxiety symptoms in a large population-based developmental cohort. Using summary statistics files from the Psychiatric Genomics Consortium Freeze 2 AN and Freeze 1 OCD GWAS, we first conducted an AN/OCD transdiagnostic GWAS meta-analysis and then calculated PGS for AN, OCD, and AN/OCD in participants from the Avon Longitudinal Study of Parents and Children with available genetic and phenotype data on eating disorder, OCD, and anxiety diagnoses and symptoms (sample size 3,212–5,369 per phenotype). We observed sex differences in the PGS prediction of eating disorder, OCD, and anxiety-related phenotypes, with AN genetic risk manifesting at an earlier age and playing a more prominent role in eating disorder phenotypes in boys than in girls. Compulsive exercise was the only phenotype predicted by all three PGS (e.g.,*P*_*AN(boys)*_ = 0.0141 at age 14; *P*_*OCD(girls)*_ = 0.0070 at age 16; *P*_*AN/OCD(all)*_ = 0.0297 at age 14). Our results suggest that earlier detection of eating disorder, OCD, and anxiety-related symptoms could be made possible by including measurement of genetic risk for these psychiatric conditions while being mindful of sex differences.

## INTRODUCTION

Eating disorders and obsessive-compulsive disorder (OCD) are serious psychiatric conditions with high social, psychological, and physical impact.^1-3^ Clinical, epidemiological, and genetic findings support an overlap between eating disorders, particularly anorexia nervosa (AN), and OCD.^4-15^ Whilst research on eating disorders and OCD comorbidity has primarily focused on diagnoses, many symptoms and behaviors are common to both diagnoses, spanning diagnostic categories, and their presence often precedes disorder onset.^16,17^ Little research has examined these associations—or symptom phenotypes—in a developmental context. Premorbid OCD symptoms, and anxiety disorders or symptoms are common in patients with AN.^18^ Childhood anxiety may precede eating disorder symptoms and AN in adolescence,^19^ and shared genetic and environmental influences play a role in anxiety and disordered eating symptoms.^20^ Though no longer classified as an anxiety disorder,^1^ OCD is highly comorbid with anxiety disorders and includes anxiety symptoms,^21^ especially in children.^22^ An improved understanding of the overlap among eating disorders and OCD, and intermediate phenotypes such as anxiety symptoms could aid in conceptualizing mechanisms and processes contributing to the clinical and genetic overlap between these disorders. Additionally, symptom dimensions may transmute over development, shifting from childhood obsessive-compulsive symptoms to adolescent eating disorders^23,24^ and vice versa. Thus, shared and unique risk factors may contribute to symptoms of OCD and eating disorders across development.

Genome-wide association studies (GWAS) of AN^14^ and OCD^25^ have provided important insights into the highly polygenic architecture of these disorders and their positive genetic correlation.^14^ Application of polygenic scores (PGS)—the weighted sum of common risk variants per individual—examine the genetic architecture of complex traits using evidence for association from variants below the stringent threshold for genome-wide significance.^26^ The use of PGS has been validated across psychiatric diagnoses and symptom-level measures,^27-33^ demonstrating that genetic variants associated with risk are often shared across diagnostic categories.^33^ Moreover, transdiagnostic PGS (determined by either AN or OCD case status) of genetically correlated disorders may enhance predictive power for either disorder.^34^

Sex differences in the prevalence and presentation of eating disorders, anxiety disorders, and OCD warrant sex-specific examination of risk factors. While the majority of AN cases are female,^35,36^ AN in males often has an earlier age of onset and is likely to be more severe.^37-39^ Similarly, the lifetime prevalence of eating disorders is much higher in females than males,^35,36^ possibly with the exception of subthreshold binge eating.^35^ The lifetime prevalence of anxiety disorders is up to 60% higher in women than in men.^40^ In the case of OCD, childhood onset is more common among males and adolescent onset is more common among females.^41^ Importantly, sex differences in the presentation of symptoms such as restraint and weight and shape concern in eating disorders^37^ and contamination/cleaning and sexual/religious symptoms in OCD^42^ have also been reported. Given these discrepancies, we could expect: a) notable sex differences in the role of genetic risk and eating disorders, OCD, and anxiety symptom phenotypes; and b) that genetic risk may be more impactful and predictive for boys, especially in the case of eating disorders.

This study examined whether the AN, OCD, or AN/OCD PGS predicts eating disorders, OCD, and anxiety symptom dimensions or diagnoses using a developmental framework in male and female participants from a population-based cohort. Our main hypothesis was that AN/OCD PGS would demonstrate better statistical power than AN or OCD PGS, and the transdiagnostic PGS would evidence the most benefit compared with single-trait PGS when predicting intermediate phenotypes *shared* across the two disorders, such as generalized anxiety or worrying.

We also hypothesized that symptom dimensions specific to each disorder would be predicted by disorder-specific PGS (e.g., thin ideal internalization by AN, or symmetry/checking behavior by OCD).^43^ Finally, in light of the differences in lifetime prevalence and/or age of onset of eating disorders, OCD, and anxiety disorders between sexes, we hypothesized that there would be significant sex differences in whether AN, OCD, and AN/OCD PGS predicted eating disorders and anxiety phenotypes, and high AN genetic risk would play a larger role in predicting eating disorder symptoms in boys than girls.

## MATERIALS AND METHODS

### Participants

The Avon Longitudinal Study of Parents and Children (ALSPAC) is a longitudinal, population-based study of women and their children.^44^ All pregnant women living in Avon, United Kingdom who were expected to deliver between April 1, 1991, and December 31, 1992 were invited to participate. Children from 14,541 pregnancies were enrolled, 13988 of whom were alive at one year. An additional 713 children were enrolled at or after age 7.^44^ Please note that the study website contains details of all the data that is available through a fully searchable data dictionary and variable search tool (http://www.bristol.ac.uk/alspac/researchers/our-data/). Ethical approval was obtained from the ALSPAC Ethics and Law Committee and the Local Research Ethics Committees.

For genetic analyses, we used post-quality control (QC) dosage files for 7,977 unrelated participants,^45,46^ 7,779 of whom passed additional QC performed as a part of this study (***Supplementary Information***). The final number of participants with phenotype and genotype data was 3,212–5,369 (see ***Tables 2–4*** for phenotype-specific sample sizes).

### Measures

***Tables 1*** provides a list of all measures, assessment timepoints, and methods of administration.

**Table 1.**
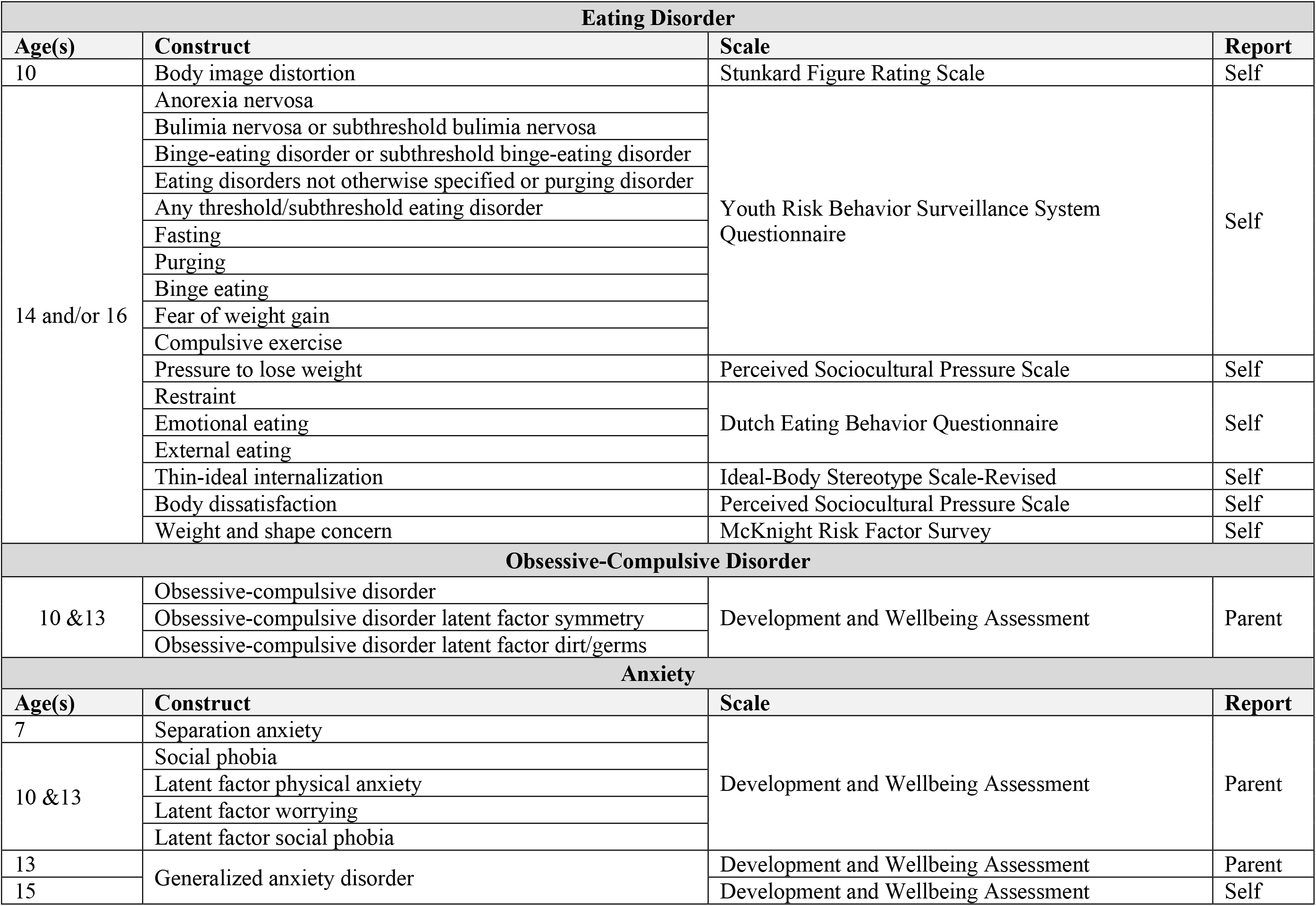
Eating disorder, obsessive-compulsive disorder, and anxiety diagnostic and symptom-based constructs.

**Table 2.** Prediction of eating disorder, obsessive-compulsive disorder, and anxiety symptom dimensions and diagnoses using polygenic scores in all participants^a^.

| Eating Disorder Symptom Dimensions and Diagnoses |  |  |  |  |  |  |  |
| --- | --- | --- | --- | --- | --- | --- | --- |
| Phenotype | Sample size | PGS | $P_T$ | N SNPs | $R^2$ | $\beta$ (SE) | Empirical $P$ |
| Body image distortion at age 10 | 4758 | AN | 0.4 | 57861 | 0.0002 | -0.0084 (0.0098) | 0.7889 |
|  |  | OCD | 0.2 | 38302 | 0.0004 | -0.0139 (0.0097) | 0.4322 |
|  |  | AN/OCD | 0.05 | 18540 | 0.0001 | -0.0069 (0.0098) | 0.8724 |
| AN at age 14<sup>b</sup> | 106 cases,<br>4129 controls | AN | 0.05 | 15063 | 0.0044 | 0.1948 (0.0986) | 0.1545 |
|  |  | OCD | 0.001 | 601 | 0.0019 | 0.1271 (0.0979) | 0.5193 |
|  |  | AN/OCD | 0.3 | 54054 | 0.0017 | 0.1223 (0.0992) | 0.5160 |
| Bulimia nervosa or subthreshold bulimia nervosa at age 14<sup>b</sup> | 63 cases,<br>4172 controls | AN | 0.001 | 1081 | 0.0012 | 0.1080 (0.1257) | 0.7847 |
|  |  | OCD | 0.001 | 601 | 0.0012 | -0.1095 (0.1279) | 0.8268 |
|  |  | AN/OCD | 0.001 | 1388 | 0.0026 | 0.1596 (0.1272) | 0.5025 |
| Binge-eating disorder or subthreshold binge-eating disorder at age 14<sup>b,c</sup> | 25 cases,<br>4210 controls | AN | -- | -- | -- | -- | -- |
|  |  | OCD | -- | -- | -- | -- | -- |
|  |  | AN/OCD | -- | -- | -- | -- | -- |
| Eating disorders not otherwise specified or purging disorder at age 14<sup>b</sup> | 593 cases,<br>3642 controls | AN | 0.01 | 5029 | 0.0012 | 0.0755 (0.0445) | 0.2588 |
|  |  | OCD | 0.001 | 601 | 0.0004 | -0.0418 (0.0444) | 0.7729 |
|  |  | AN/OCD | 0.05 | 18540 | 0.0010 | 0.0694 (0.0445) | 0.3181 |
| Any threshold/subthreshold eating disorder at age 14<sup>b</sup> | 787 cases,<br>3448 controls | AN | 0.01 | 5029 | 0.0015 | 0.0780 (0.0396) | 0.1471 |
|  |  | OCD | 0.01 | 3915 | 0.0001 | -0.0229 (0.0396) | 0.9556 |
|  |  | AN/OCD | 0.01 | 6379 | 0.0013 | 0.0737 (0.0398) | 0.1775 |
| Fasting at age 14<sup>b</sup> | 271 cases,<br>3861 controls | AN | 0.3 | 48847 | 0.0006 | 0.0633 (0.0641) | 0.7006 |
|  |  | OCD | 0.4 | 60239 | 0.0007 | -0.0653 (0.0628) | 0.7066 |
|  |  | AN/OCD | 0.5 | 68009 | 0.0009 | 0.0761 (0.0640) | 0.5449 |
| Purging at age 14 | 4132 | AN | 0.05 | 15063 | 0.0006 | 0.0063 (0.0040) | 0.3280 |
|  |  | OCD | 0.001 | 601 | 0.0008 | -0.0071 (0.0040) | 0.2441 |
|  |  | AN/OCD | 0.1 | 28747 | 0.0007 | 0.0070 (0.0040) | 0.2333 |
| Binge eating at age 14 | 4167 | AN | 0.01 | 5029 | 0.0001 | 0.0050 (0.0064) | 0.8335 |
|  |  | OCD | 0.1 | 23472 | 0.0007 | 0.0109 (0.0064) | 0.2751 |
|  |  | AN/OCD | 0.001 | 1388 | 0.0002 | 0.0062 (0.0064) | 0.6942 |
| Fear of weight gain at age 14 | 4127 | AN | 0.01 | 5029 | 0.0003 | 0.0128 (0.0107) | 0.5470 |
|  |  | OCD | 0.05 | 13919 | 0.0002 | 0.0092 (0.0107) | 0.8251 |
|  |  | AN/OCD | 1 | 82076 | 0.0002 | 0.0087 (0.0108) | 0.8113 |
| Compulsive exercise at age 14 | 4027 | AN | 0.05 | 15063 | 0.0015 | 0.0247 (0.0099) | **0.0448\*** |
|  |  | OCD | 0.001 | 601 | 0.0004 | -0.0131 (0.0099) | 0.5130 |
|  |  | AN/OCD | 0.01 | 6379 | 0.0018 | 0.0265 (0.0100) | **0.0297\*** |
| Pressure to lose weight at age 14 | 4116 | AN | 0.001 | 1081 | 0.0009 | 0.0524 (0.0279) | 0.1802 |
|  |  | OCD | 0.001 | 601 | 0.0007 | -0.0494 (0.0281) | 0.2460 |
|  |  | AN/OCD | 0.001 | 1388 | 0.0010 | 0.0573 (0.0283) | 0.1322 |
| Restraint at age 14 | 4074 | AN | 0.3 | 48847 | 0.0014 | 0.0434 (0.0179) | 0.0557 |
|  |  | OCD | 0.05 | 13919 | 0.0002 | 0.0140 (0.0178) | 0.8611 |
|  |  | AN/OCD | 0.2 | 43351 | 0.0020 | 0.0518 (0.0179) | **0.0147\*** |
| Emotional eating at age 14 | 3927 | AN | 0.001 | 1081 | 0.0001 | 0.0515 (0.0888) | 0.9393 |
|  |  | OCD | 0.001 | 601 | 0.0004 | -0.1166 (0.0890) | 0.5123 |
|  |  | AN/OCD | 0.05 | 18540 | 0.0001 | 0.0552 (0.0893) | 0.9126 |
| External eating at age 14 | 3608 | AN | 0.001 | 1081 | 0.0001 | 0.0403 (0.0551) | 0.8617 |
|  |  | OCD | 0.05 | 13919 | 0.0005 | 0.0775 (0.0554) | 0.4512 |
|  |  | AN/OCD | 0.001 | 1388 | 0.0003 | 0.0593 (0.0558) | 0.6353 |
| Thin ideal internalization at age 14 | 4053 | AN | 0.001 | 1081 | 0.0010 | 0.0864 (0.0424) | 0.1351 |
|  |  | OCD | 0.1 | 23472 | 0.0008 | 0.0784 (0.0426) | 0.2164 |
|  |  | AN/OCD | 0.001 | 1388 | 0.0006 | 0.0659 (0.0430) | 0.3325 |
| Body dissatisfaction at age 14 | 4169 | AN | 0.1 | 24182 | 0.0015 | 0.3044 (0.1204) | **0.0410\*** |
|  |  | OCD | 0.001 | 601 | 0.0003 | −0.1294 (0.1202) | 0.6720 |
|  |  | AN/OCD | 0.2 | 43351 | 0.0018 | 0.3329 (0.1209) | **0.0200\*** |
| Weight and shape concern at age 14 | 4164 | AN | 0.001 | 1081 | 0.0002 | 0.0269 (0.0284) | 0.7268 |
|  |  | OCD | 0.05 | 13919 | 0.0002 | 0.0248 (0.0285) | 0.8105 |
|  |  | AN/OCD | 1 | 82076 | 0.0002 | 0.0268 (0.0289) | 0.7250 |
| AN at age 16<sup>b</sup> | 71 cases,<br>3511 controls | AN | 0.001 | 1081 | 0.0060 | −0.2354 (0.1213) | 0.1643 |
|  |  | OCD | 0.01 | 3915 | 0.0008 | 0.0824 (0.1182) | 0.9028 |
|  |  | AN/OCD | 0.1 | 28747 | 0.0037 | −0.1825 (0.1189) | 0.3203 |
| Bulimia nervosa at age 16<sup>b</sup> | 146 cases,<br>3436 controls | AN | 0.001 | 1081 | 0.0004 | 0.0555 (0.0845) | 0.9059 |
|  |  | OCD | 0.1 | 23472 | 0.0005 | 0.0629 (0.0843) | 0.8835 |
|  |  | AN/OCD | 0.001 | 1388 | 0.0017 | 0.1120 (0.0839) | 0.4432 |
| Binge-eating disorder at age 16<sup>b</sup> | 58 cases,<br>3524 controls | AN | 0.3 | 48847 | 0.0026 | 0.1583 (0.1336) | 0.5600 |
|  |  | OCD | 0.1 | 23472 | 0.0033 | 0.1779 (0.1320) | 0.4711 |
|  |  | AN/OCD | 0.001 | 1388 | 0.0034 | 0.1806 (0.1315) | 0.4135 |
| Eating disorders not otherwise specified or purging disorder at age 16<sup>b</sup> | 1079 cases,<br>2503 controls | AN | 0.001 | 1081 | 0.0004 | −0.0370 (0.0365) | 0.6728 |
|  |  | OCD | 0.05 | 13919 | 0.0026 | 0.0935 (0.0366) | **0.0410\*** |
|  |  | AN/OCD | 0.05 | 18540 | 0.0001 | −0.0157 (0.0363) | 0.9776 |
| Any threshold/subthreshold eating disorder at age 16<sup>b</sup> | 1354 cases,<br>2228 controls | AN | 0.001 | 1081 | 0.0005 | −0.0384 (0.0345) | 0.6112 |
|  |  | OCD | 0.1 | 23472 | 0.0035 | 0.1045 (0.0345) | **0.0111\*** |
|  |  | AN/OCD | 0.2 | 43351 | < 0.0001 | 0.0068 (0.0343) | 0.9993 |
| Fasting at age 16 | 3379 | AN | 0.001 | 1081 | 0.0002 | −0.0101 (0.0114) | 0.7675 |
|  |  | OCD | 0.05 | 13919 | 0.0014 | 0.0243 (0.0112) | 0.1070 |
|  |  | AN/OCD | 1 | 82076 | 0.0001 | −0.0075 (0.0113) | 0.8968 |
| Purging at age 16 | 3402 | AN | 0.1 | 24182 | 0.0008 | 0.0142 (0.0085) | 0.2536 |
|  |  | OCD | 0.01 | 3915 | 0.0007 | 0.0125 (0.0084) | 0.3789 |
|  |  | AN/OCD | 0.05 | 18540 | 0.0012 | 0.0176 (0.0084) | 0.1045 |
| Binge eating at age 16 | 2929 | AN | 0.3 | 48847 | 0.0002 | 0.0102 (0.0122) | 0.7994 |
|  |  | OCD | 0.1 | 23472 | 0.0011 | 0.0219 (0.0120) | 0.2197 |
|  |  | AN/OCD | 0.001 | 1388 | 0.0012 | 0.0230 (0.0122) | 0.1771 |
| Compulsive exercise at age 16 | 3186 | AN | 0.1 | 24182 | 0.0002 | 0.0147 (0.0172) | 0.7860 |
|  |  | OCD | 0.05 | 13919 | 0.0034 | 0.0555 (0.0170) | **0.0042\*** |
|  |  | AN/OCD | 0.4 | 61888 | 0.0006 | 0.0227 (0.0171) | 0.4465 |
| Obsessive-Compulsive Disorder Symptom Dimensions and Diagnosis |  |  |  |  |  |  |  |
| Phenotype | Sample size | PGS | *P*<sub>T</sub> | N SNPs | *R*<sup>2</sup> | *β* (*SE*) | Empirical *P* |
| OCD at age 10<sup>b,c</sup> | 18 cases,<br>5198 controls | AN | -- | -- | -- | -- | -- |
|  |  | OCD | -- | -- | -- | -- | -- |
|  |  | AN/OCD | -- | -- | -- | -- | -- |
| OCD latent factor – symmetry, checking at age 10 | 5197 | AN | 0.001 | 1081 | 0.0009 | 0.0151 (0.0069) | 0.0949 |
|  |  | OCD | 0.2 | 38302 | 0.0001 | −0.0060 (0.0070) | 0.8131 |
|  |  | AN/OCD | 0.2 | 43351 | 0.0007 | 0.0132 (0.0070) | 0.1781 |
| OCD latent factor – dirt/germs at age 10 | 5197 | AN | 0.001 | 1081 | 0.0013 | 0.0157 (0.0060) | **0.0319\*** |
|  |  | OCD | 0.05 | 13919 | 0.0001 | 0.0032 (0.0060) | 0.9645 |
|  |  | AN/OCD | 0.2 | 43351 | 0.0002 | 0.0068 (0.0060) | 0.5745 |
| OCD at age 13<sup>b,c</sup> | 10 cases<br>4841 controls | AN | -- | -- | -- | -- | -- |
|  |  | OCD | -- | -- | -- | -- | -- |
|  |  | AN/OCD | -- | -- | -- | -- | -- |
| OCD latent factor – symmetry, checking at age 13 | 4832 | AN | 1 | 85364 | 0.0003 | 0.0125 (0.0109) | 0.5815 |
|  |  | OCD | 0.001 | 601 | 0.0002 | −0.0106 (0.0107) | 0.7313 |
|  |  | AN/OCD | 0.01 | 6379 | < 0.0001 | 0.0052 (0.0109) | 0.9669 |
| OCD latent factor – dirt/germs at age 13 | 4832 | AN | 0.05 | 15063 | 0.0001 | −0.0064 (0.0093) | 0.8866 |
|  |  | OCD | 0.05 | 13919 | 0.0009 | −0.0196 (0.0092) | 0.1208 |
|  |  | AN/OCD | 0.001 | 1388 | < 0.0001 | −0.0045 (0.0093) | 0.9646 |
| Anxiety Symptom Dimensions and Diagnoses |  |  |  |  |  |  |  |
| Phenotype | Sample size | PGS | P<sub>T</sub> | N SNPs | R<sup>2</sup> | β (SE) | Empirical P |
| Separation anxiety at age 7<sup>b</sup> | 148 cases,<br>5214 controls | AN | 0.3 | 48847 | 0.0072 | 0.2475 (0.0841) | **0.0132\*** |
|  |  | OCD | 0.05 | 13919 | 0.0001 | 0.0308 (0.0831) | 0.9954 |
|  |  | AN/OCD | 0.2 | 43351 | 0.0039 | 0.1812 (0.0843) | 0.0982 |
| Specific phobia at age 7<sup>b</sup> | 104 cases,<br>5265 controls | AN | 0.01 | 5029 | 0.0063 | 0.2432 (0.1001) | 0.0501 |
|  |  | OCD | 0.001 | 601 | 0.0010 | −0.0955 (0.0988) | 0.7517 |
|  |  | AN/OCD | 0.01 | 6379 | 0.0085 | 0.2809 (0.0999) | **0.0181\*** |
| Latent factor – physical anxiety at age 10 | 5197 | AN | 0.01 | 5029 | 0.0009 | 0.0221 (0.0102) | 0.0945 |
|  |  | OCD | 0.2 | 38302 | 0.0001 | −0.0065 (0.0102) | 0.9302 |
|  |  | AN/OCD | 0.1 | 28747 | 0.0007 | 0.0195 (0.0102) | 0.1675 |
| Latent factor – worrying at age 10 | 5197 | AN | 0.01 | 5029 | 0.0016 | 0.0283 (0.0099) | **0.0163\*** |
|  |  | OCD | 0.1 | 23472 | 0.0003 | 0.0131 (0.0099) | 0.5107 |
|  |  | AN/OCD | 0.1 | 28747 | 0.0004 | 0.0150 (0.0099) | 0.3376 |
| Latent factor – social phobia at age 10 | 5197 | AN | 0.001 | 1081 | 0.0005 | 0.0140 (0.0085) | 0.2810 |
|  |  | OCD | 0.01 | 3915 | 0.0009 | −0.0184 (0.0085) | 0.1102 |
|  |  | AN/OCD | 0.01 | 6379 | 0.0002 | 0.0083 (0.0086) | 0.7001 |
| Social phobia at age 13<sup>b</sup> | 55 cases,<br>4795 controls | AN | 0.3 | 48847 | 0.0015 | 0.1254 (0.1366) | 0.7521 |
|  |  | OCD | 0.01 | 3915 | 0.0088 | 0.3036 (0.1359) | 0.0959 |
|  |  | AN/OCD | 0.001 | 1388 | 0.0015 | 0.1258 (0.1371) | 0.7412 |
| Generalized anxiety disorder at age 13<sup>b</sup> | 154 cases,<br>4687 controls | AN | 0.01 | 5029 | 0.0029 | 0.1544 (0.0826) | 0.1916 |
|  |  | OCD | 1 | 91297 | 0.0019 | 0.1248 (0.0823) | 0.3813 |
|  |  | AN/OCD | 0.01 | 6379 | 0.0025 | 0.1422 (0.0830) | 0.2363 |
| Latent factor – physical anxiety at age 13 | 4832 | AN | 1 | 85364 | 0.0003 | −0.0118 (0.0096) | 0.5315 |
|  |  | OCD | 0.01 | 3915 | 0.0001 | −0.0067 (0.0095) | 0.9022 |
|  |  | AN/OCD | 0.2 | 43351 | 0.0002 | −0.0098 (0.0096) | 0.6668 |
| Latent factor – worrying at age 13 | 4832 | AN | 0.3 | 48847 | 0.0006 | 0.0197 (0.0114) | 0.2414 |
|  |  | OCD | 0.001 | 601 | 0.0003 | −0.0131 (0.0112) | 0.6147 |
|  |  | AN/OCD | 0.2 | 43351 | 0.0003 | 0.0134 (0.0114) | 0.5571 |
| Latent factor – social phobia at age 13 | 4832 | AN | 0.3 | 48847 | 0.0014 | 0.0278 (0.0108) | **0.0380\*** |
|  |  | OCD | 0.001 | 601 | 0.0005 | −0.0168 (0.0107) | 0.3497 |
|  |  | AN/OCD | 0.2 | 43351 | 0.0006 | 0.0179 (0.0108) | 0.2637 |
| Generalized anxiety disorder at age 15<sup>b</sup> | 196 cases,<br>3712 controls | AN | 0.001 | 1081 | 0.0019 | 0.1136 (0.0723) | 0.3184 |
|  |  | OCD | 0.1 | 23472 | 0.0006 | 0.0657 (0.0738) | 0.8034 |
|  |  | AN/OCD | 0.001 | 1388 | 0.0003 | 0.0453 (0.0742) | 0.9199 |

<sup>a</sup> Principal components 1–5 were used as covariates.
<sup>b</sup> Binary phenotype.
<sup>c</sup> Due to insufficient statistical power, any binary measure with less than 50 cases is not included in the final analysis.
\* (also bolded) Statistically significant after $P$ for regression was corrected for multiple testing via 10,000 permutations (significance threshold=0.05).
Abbreviations: PGS=polygenic score; $P_T$ =best discovery $P$ threshold; SNP=single nucleotide polymorphism; $R^2$ =Nagelkerke's pseudo- $R^2$ for total variance explained in target phenotype; $\beta$ =standardized beta regression coefficient; $SE$ =standard error; AN=anorexia nervosa; OCD=obsessive-compulsive disorder; AN/OCD=anorexia nervosa/obsessive-compulsive transdiagnostic phenotype.

Eating disorder symptoms for the previous year were evaluated at ages 14 and 16 using questions adapted from the Youth Risk Behavior Surveillance System Questionnaire,^47^ validated in a population-based study of adolescents.^48^ Binge-eating, purging, fasting, and compulsive exercise were characterized and categorized as described previously (***Supplementary Information***).^49,50^ Eating disorder diagnoses at ages 14 and 16 were derived using DSM-5 criteria^1^ as detailed in a previous publication by our group.^19^ Eating disorder cognitions, including: body image distortion, emotional eating, external eating, body dissatisfaction, thin ideal internalization, dietary restraint, weight concern, and shape concern were assessed by validated, age appropriate self-report measurements (***Supplementary Information***).

OCD and anxiety symptoms at age 7, 10, 13, and 15 were collected using the Development and Wellbeing Assessment (DAWBA; ***Supplementary Information***).^51,52^ Probabilities of anxiety disorder diagnoses at ages 7 (specific phobia and separation anxiety), 10 (OCD), 13 (OCD, social phobia, and generalized anxiety disorder), and 15 (generalized anxiety disorder) were determined using computer-generated DAWBA band variables,^52^ which assign the probability of the participant meeting DSM-IV criteria for an anxiety disorder. However, due to a very small number of participants scoring high on these variables, we dichotomized them to capture whether the likelihood of a diagnosis was <50% versus ≥50%. We also defined five latent OCD or anxiety factors for ages 10 and 13: (1) OCD-symmetry; (2) OCD-dirt/germs; (3) physical anxiety; (4) worrying; and (5) social phobia (***Supplementary Information***).^19^

### Data Analysis

We calculated AN, OCD, and AN/OCD PGS to predict 27 eating disorder, 6 OCD, and 11 anxiety phenotypes in the ALSPAC target sample using PRSice v2.2.^53^ AN PGS was constructed using the Anorexia Nervosa Genetics Initiative & Psychiatric Genomics Consortium (PGC) Eating Disorder Working Group Freeze 2 AN GWAS,^14^ and OCD PGS was calculated using the Freeze 1 PGC OCD GWAS.^25^ AN/OCD summary statistics files were obtained from a GWAS conducted by combining these datasets (see ***Supplementary Information, Table S1***, and ***Figure S1***). We first examined how well AN, OCD, and AN/OCD PGS predicted eating disorder, OCD, and anxiety symptom phenotypes in the combined sample and then investigated sex-specific differences. Due to insufficient power, only binary phenotypes with ≥50 cases are reported. Significance threshold was set at empirical *P*<0.05 following 10,000 permutations per target phenotype. Corresponding odds ratios and 95% confidence intervals for binary phenotypes are in ***Table S2***.

## RESULTS

### Eating disorder symptom phenotypes and diagnoses

***Figures 1a*** and ***1b*** show the significant results for eating disorder phenotypes. In the combined sample, AN PGS predicted compulsive exercise (*P* = 0.0448) and body dissatisfaction at age 14 (*P* = 0.0410), whereas OCD PGS predicted eating disorder not otherwise specified/purging disorder (*P* = 0.0410), the presence of a threshold or subthreshold eating disorder (*P* = 0.0111), and compulsive exercise (*P* < 0.0042) at age 16 (***Table 2***). In addition to predicting compulsive exercise (*P* = 0.0297) and body dissatisfaction at age 14 (*P* = 0.0200), AN/OCD PGS uniquely predicted higher restraint at age 14 (*P* = 0.0147).

**Figure 1.**
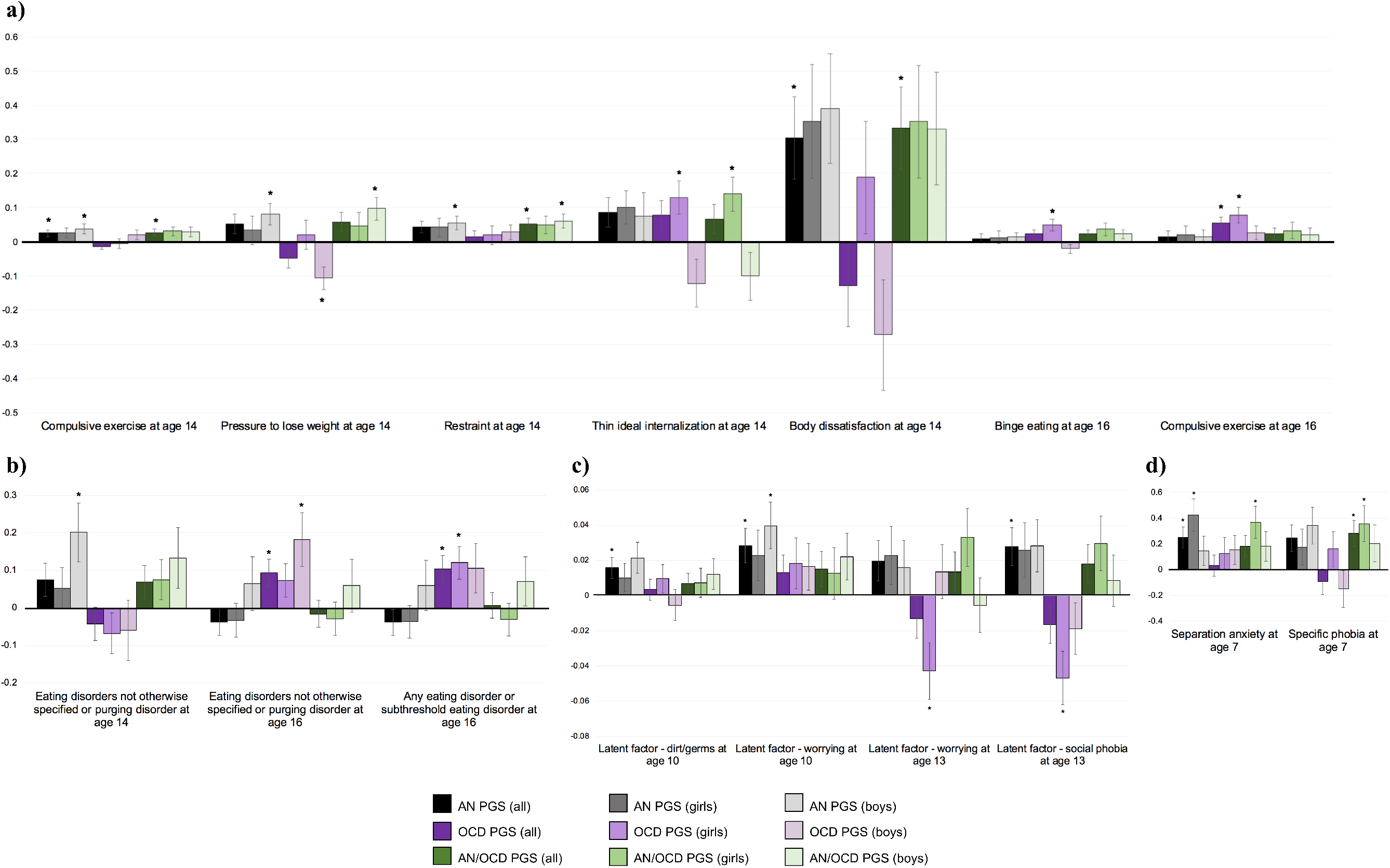
Significant results for the prediction of **(a)** eating disorder symptoms; **(b)** eating disorder diagnoses; **(c)** obsessive-compulsive disorder and anxiety-related symptoms; and **(d)** obsessive-compulsive disorder and anxiety disorder diagnoses by polygenic scores in all participants (combined and by sex) during different developmental timepoints. Significant findings (threshold set at *p*> 0.05 following 10,000 permutations per target phenotype) are indicated with an asterisk. The bars depict standardized beta regression coefficients, and the lines depict standard errors.

In girls, none of the eating disorder-related constructs were predicted by AN PGS (***Table 3***). OCD PGS predicted higher thin-ideal internalization at age 14 (*P=*0.0361) as well as presence of a threshold or subthreshold eating disorder (*P*=0.0232), binge eating (*P*=0.0264), and compulsive exercise at age 16 (*P*=0.0070). AN/OCD PGS predicted increased thin-ideal internalization at age 14 (*P*=0.0185).

In boys, AN PGS predicted eating disorder not otherwise specified/purging disorder (*P*=0.0400), increased compulsive exercise (*P*=0.0141), and greater internal pressure to lose weight (*P*=0.0432) at age 14 (***Table 3***). OCD PGS predicted less internal pressure to lose weight at age 14 (*P=*0.0051), and the presence of any eating disorder or subthreshold eating disorder at age 16 (*P*=0.0479). AN/OCD PGS predicted greater internal pressure to lose weight (*P*=0.0122) and higher restraint at age 14 (*P*=0.013).

**Table 3.** Prediction of eating disorder, obsessive-compulsive disorder, and anxiety symptom dimensions and diagnoses using polygenic scores in girls^a^.

| Eating Disorder Symptom Dimensions and Diagnoses |  |  |  |  |  |  |  |
| --- | --- | --- | --- | --- | --- | --- | --- |
| Phenotype | Sample size | PGS | $P_T$ | N SNPs | $R^2$ | $\beta$ (SE) | Empirical $P$ |
| Body image distortion at age 10 | 2512 | AN | 0.001 | 1079 | 0.0005 | -0.0158 (0.0135) | 0.5689 |
|  |  | OCD | 0.001 | 602 | 0.0006 | 0.0165 (0.0135) | 0.5702 |
|  |  | AN/OCD | 0.001 | 1385 | 0.0002 | -0.0095 (0.0136) | 0.8783 |
| AN at age 14<sup>b</sup> | 78 cases,<br>2242 controls | AN | 0.05 | 15037 | 0.0062 | 0.2254 (0.1173) | 0.1731 |
|  |  | OCD | 0.001 | 602 | 0.0009 | 0.0863 (0.1149) | 0.8775 |
|  |  | AN/OCD | 0.001 | 1385 | 0.0059 | 0.2167 (0.1163) | 0.1865 |
| Bulimia nervosa or subthreshold bulimia nervosa at age 14<sup>b,c</sup> | 44 cases, 2276 controls | AN | -- | -- | -- | -- | -- |
|  |  | OCD | -- | -- | -- | -- | -- |
|  |  | AN/OCD | -- | -- | -- | -- | -- |
| Binge-eating disorder or subthreshold binge-eating disorder at age 14<sup>b,c</sup> | 17 cases, 2303 controls | AN | -- | -- | -- | -- | -- |
|  |  | OCD | -- | -- | -- | -- | -- |
|  |  | AN/OCD | -- | -- | -- | -- | -- |
| Eating disorders not otherwise specified or purging disorder at age 14<sup>b</sup> | 419 cases, 1901 controls | AN | 0.3 | 48846 | 0.0007 | 0.0531 (0.0543) | 0.7027 |
|  |  | OCD | 0.01 | 3917 | 0.0011 | -0.0680 (0.0545) | 0.5441 |
|  |  | AN/OCD | 0.4 | 61950 | 0.0014 | 0.0754 (0.0542) | 0.4122 |
| Any threshold/subthreshold eating disorder at age 14<sup>b</sup> | 558 cases, 1762 controls | AN | 0.05 | 15037 | 0.0006 | 0.0490 (0.0492) | 0.6948 |
|  |  | OCD | 0.01 | 3917 | 0.0002 | -0.0294 (0.0486) | 0.9452 |
|  |  | AN/OCD | 0.3 | 54071 | 0.0017 | 0.0801 (0.0488) | 0.2844 |
| Fasting at age 14<sup>b</sup> | 227 cases, 2047 controls | AN | 1 | 85306 | 0.0015 | 0.0890 (0.0706) | 0.4959 |
|  |  | OCD | 0.5 | 68461 | 0.0005 | -0.0533 (0.0697) | 0.8726 |
|  |  | AN/OCD | 0.5 | 68072 | 0.0024 | 0.1145 (0.0709) | 0.2870 |
| Purging at age 14 | 2278 | AN | 0.4 | 57808 | 0.0013 | 0.0116 (0.0067) | 0.2441 |
|  |  | OCD | 0.001 | 602 | 0.0011 | -0.0106 (0.0067) | 0.3376 |
|  |  | AN/OCD | 0.05 | 18537 | 0.0016 | 0.0129 (0.0067) | 0.1606 |
| Binge eating at age 14 | 2289 | AN | 0.05 | 15037 | 0.0008 | -0.0124 (0.0094) | 0.4818 |
|  |  | OCD | 0.1 | 23463 | 0.0010 | 0.0143 (0.0093) | 0.3599 |
|  |  | AN/OCD | 0.05 | 18537 | 0.0004 | -0.0095 (0.0094) | 0.6817 |
| Fear of weight gain at age 14 | 2277 | AN | 0.4 | 57808 | 0.0003 | 0.0124 (0.0161) | 0.8368 |
|  |  | OCD | 0.001 | 602 | 0.0011 | 0.0251 (0.0161) | 0.3431 |
|  |  | AN/OCD | 0.5 | 68072 | 0.0003 | 0.0134 (0.0161) | 0.7958 |
| Compulsive exercise at age 14 | 2240 | AN | 0.05 | 15037 | 0.0016 | 0.0266 (0.0142) | 0.1809 |
|  |  | OCD | 0.001 | 602 | 0.0001 | -0.0053 (0.0141) | 0.9943 |
|  |  | AN/OCD | 0.01 | 6370 | 0.0021 | 0.0306 (0.0141) | 0.0955 |
| Pressure to lose weight at age 14 | 2263 | AN | 0.001 | 1079 | 0.0003 | 0.0334 (0.0420) | 0.8185 |
|  |  | OCD | 0.3 | 50361 | 0.0001 | 0.0205 (0.0422) | 0.9768 |
|  |  | AN/OCD | 0.001 | 1385 | 0.0005 | 0.0454 (0.0422) | 0.6297 |
| Restraint at age 14 | 2257 | AN | 0.3 | 48846 | 0.0011 | 0.0423 (0.0265) | 0.3096 |
|  |  | OCD | 0.001 | 602 | 0.0002 | 0.0194 (0.0263) | 0.8930 |
|  |  | AN/OCD | 0.1 | 28752 | 0.0015 | 0.0489 (0.0265) | 0.1852 |
| Emotional eating at age 14 | 2167 | AN | 0.4 | 57808 | 0.0006 | -0.1482 (0.1276) | 0.5808 |
|  |  | OCD | 0.3 | 50361 | 0.0015 | 0.2301 (0.1261) | 0.2207 |
|  |  | AN/OCD | 0.4 | 61950 | 0.0005 | -0.1370 (0.1276) | 0.6266 |
| External eating at age 14 | 2068 | AN | 0.05 | 15037 | 0.0008 | −0.0926 (0.0722) | 0.4879 |
|  |  | OCD | 0.1 | 23463 | 0.0007 | 0.0848 (0.0711) | 0.5969 |
|  |  | AN/OCD | 0.2 | 43327 | 0.0001 | 0.0277 (0.0724) | 0.9895 |
| Thin ideal internalization at age 14 | 2283 | AN | 0.001 | 1079 | 0.0018 | 0.1010 (0.0495) | 0.1303 |
|  |  | OCD | 0.1 | 23463 | 0.0030 | 0.1287 (0.0492) | **0.0361<sup>+</sup>** |
|  |  | AN/OCD | 0.001 | 1385 | 0.0034 | 0.1394 (0.0497) | **0.0185<sup>+</sup>** |
| Body dissatisfaction at age 14 | 2297 | AN | 0.2 | 37940 | 0.0020 | 0.3528 (0.1654) | 0.1050 |
|  |  | OCD | 0.05 | 13923 | 0.0006 | 0.1886 (0.1640) | 0.6161 |
|  |  | AN/OCD | 0.2 | 43327 | 0.0020 | 0.3521 (0.1650) | 0.1015 |
| Weight and shape concern at age 14 | 2299 | AN | 0.1 | 24166 | 0.0001 | −0.0233 (0.0412) | 0.9453 |
|  |  | OCD | 0.001 | 602 | 0.0008 | 0.0562 (0.0410) | 0.4725 |
|  |  | AN/OCD | 1 | 82082 | 0.0001 | 0.0216 (0.0409) | 0.9551 |
| AN at age 16<sup>b</sup> | 56 cases,<br>2035 controls | AN | 0.001 | 1079 | 0.0021 | −0.1342 (0.1380) | 0.6993 |
|  |  | OCD | 0.05 | 13923 | 0.0017 | 0.1183 (0.1356) | 0.8054 |
|  |  | AN/OCD | 1 | 82082 | 0.0020 | −0.1295 (0.1348) | 0.7006 |
| Bulimia nervosa at age 16<sup>b</sup> | 121 cases,<br>1970 controls | AN | 0.01 | 5029 | 0.0012 | −0.0908 (0.0943) | 0.7122 |
|  |  | OCD | 0.1 | 23463 | 0.0017 | 0.1057 (0.0930) | 0.6295 |
|  |  | AN/OCD | 0.001 | 1385 | 0.0018 | 0.1089 (0.0932) | 0.5619 |
| Binge-eating disorder at age 16<sup>b,c</sup> | 48 cases,<br>2043 controls | AN | -- | -- | -- | -- | -- |
|  |  | OCD | -- | -- | -- | -- | -- |
|  |  | AN/OCD | -- | -- | -- | -- | -- |
| Eating disorders not otherwise specified or purging disorder at age 16<sup>b</sup> | 843 cases,<br>1248 controls | AN | 0.001 | 1079 | 0.0003 | −0.0326 (0.0449) | 0.8643 |
|  |  | OCD | 0.05 | 13923 | 0.0018 | 0.0736 (0.0447) | 0.3027 |
|  |  | AN/OCD | 0.01 | 6370 | 0.0003 | −0.0288 (0.0448) | 0.9038 |
| Any threshold/subthreshold eating disorder at age 16<sup>b</sup> | 1068 cases,<br>1023 controls | AN | 0.001 | 1079 | 0.0005 | −0.0369 (0.0439) | 0.7914 |
|  |  | OCD | 0.1 | 23463 | 0.0049 | 0.1206 (0.0434) | **0.0232<sup>+</sup>** |
|  |  | AN/OCD | 0.3 | 54071 | 0.0003 | −0.0306 (0.0431) | 0.8700 |
| Fasting at age 16 | 1972 | AN | 0.2 | 37940 | 0.0005 | −0.0181 (0.0180) | 0.6891 |
|  |  | OCD | 0.05 | 13923 | 0.0025 | 0.0395 (0.0178) | 0.0951 |
|  |  | AN/OCD | 1 | 82082 | 0.0004 | −0.0158 (0.0178) | 0.7573 |
| Purging at age 16 | 1990 | AN | 0.1 | 24166 | 0.0010 | 0.0196 (0.0139) | 0.4144 |
|  |  | OCD | 0.001 | 602 | 0.0009 | 0.0180 (0.0137) | 0.5050 |
|  |  | AN/OCD | 0.05 | 18537 | 0.0021 | 0.0281 (0.0138) | 0.1312 |
| Binge eating at age 16 | 1715 | AN | 0.3 | 48846 | 0.0003 | 0.0129 (0.0184) | 0.8860 |
|  |  | OCD | 0.1 | 23463 | 0.0043 | 0.0489 (0.0181) | **0.0264<sup>+</sup>** |
|  |  | AN/OCD | 0.001 | 1385 | 0.0023 | 0.0362 (0.0183) | 0.1468 |
| Compulsive exercise at age 16 | 1840 | AN | 1 | 85306 | 0.0004 | 0.0210 (0.0245) | 0.7928 |
|  |  | OCD | 0.1 | 23463 | 0.0056 | 0.0776 (0.0241) | **0.0070<sup>+</sup>** |
|  |  | AN/OCD | 0.4 | 61950 | 0.0010 | 0.0327 (0.0244) | 0.4451 |
| Obsessive-Compulsive Disorder Symptom Dimensions and Diagnosis |  |  |  |  |  |  |  |
| Phenotype | Sample size | PGS | P<sub>T</sub> | N SNPs | R<sup>2</sup> | β (SE) | Empirical P |
| OCD at age 10<sup>b,c</sup> | < 5 cases | AN | -- | -- | -- | -- | -- |
|  |  | OCD | -- | -- | -- | -- | -- |
|  |  | AN/OCD | -- | -- | -- | -- | -- |
| OCD latent factor – symmetry, checking at age 10 | 2590 | AN | 0.01 | 5029 | 0.0008 | 0.0136 (0.0092) | 0.3740 |
|  |  | OCD | 0.05 | 13923 | 0.0006 | 0.0118 (0.0092) | 0.5210 |
|  |  | AN/OCD | 0.2 | 43327 | 0.0007 | 0.0125 (0.0093) | 0.4549 |
| OCD latent factor – dirt/germs at age 10 | 2590 | AN | 0.001 | 1079 | 0.0006 | 0.0099 (0.0081) | 0.5269 |
|  |  | OCD | 0.05 | 13923 | 0.0005 | 0.0094 (0.0081) | 0.6212 |
|  |  | AN/OCD | 0.2 | 43327 | 0.0003 | 0.0071 (0.0082) | 0.7768 |
| OCD at age 13<sup>b,c</sup> | < 5 cases | AN | -- | -- | -- | -- | -- |
|  |  | OCD | -- | -- | -- | -- | -- |
|  |  | AN/OCD | -- | -- | -- | -- | -- |
| OCD latent factor – symmetry, checking at age 13 | 2421 | AN | 0.05 | 15037 | 0.0011 | −0.0250 (0.0150) | 0.2739 |
|  |  | OCD | 0.1 | 23463 | 0.0007 | 0.0196 (0.0148) | 0.4988 |
|  |  | AN/OCD | 0.01 | 6370 | 0.0003 | −0.0127 (0.0151) | 0.7972 |
| OCD latent factor – dirt/germs at age 13 | 2421 | AN | 1 | 85306 | 0.0006 | 0.0160 (0.0128) | 0.5192 |
|  |  | OCD | 1 | 91289 | 0.0012 | −0.0216 (0.0127) | 0.2652 |
|  |  | AN/OCD | 1 | 82082 | 0.0006 | 0.0161 (0.0129) | 0.5044 |
| Anxiety Symptom Dimensions and Diagnoses |  |  |  |  |  |  |  |
| Phenotype | Sample size | PGS | $P_T$ | N SNPs | $R^2$ | $\beta$ (SE) | Empirical $P$ |
| Separation anxiety at age 7<sup>b</sup> | 68 cases,<br>2544 controls | AN | 0.2 | 37940 | 0.0203 | 0.4229 (0.1256) | **0.0035** * |
|  |  | OCD | 1 | 91289 | 0.0018 | 0.1243 (0.1226) | 0.7173 |
|  |  | AN/OCD | 0.1 | 28752 | 0.0153 | 0.3655 (0.1251) | **0.0134** * |
| Specific phobia at age 7<sup>b</sup> | 53 cases,<br>2564 controls | AN | 0.01 | 5029 | 0.0033 | 0.1746 (0.1396) | 0.5141 |
|  |  | OCD | 1 | 91289 | 0.0028 | 0.1590 (0.1374) | 0.6183 |
|  |  | AN/OCD | 0.01 | 6370 | 0.0138 | 0.3562 (0.1401) | **0.0378** * |
| Latent factor – physical anxiety at age 10 | 2590 | AN | 0.001 | 1079 | 0.0010 | 0.0234 (0.0143) | 0.2819 |
|  |  | OCD | 0.001 | 602 | 0.0012 | −0.0254 (0.0145) | 0.2500 |
|  |  | AN/OCD | 0.3 | 54071 | 0.0009 | 0.0227 (0.0145) | 0.3147 |
| Latent factor – worrying at age 10 | 2590 | AN | 0.001 | 1079 | 0.0010 | 0.0228 (0.0144) | 0.3139 |
|  |  | OCD | 0.05 | 13923 | 0.0006 | 0.0181 (0.0145) | 0.5495 |
|  |  | AN/OCD | 1 | 82082 | 0.0003 | 0.0125 (0.0147) | 0.7854 |
| Latent factor – social phobia at age 10 | 2590 | AN | 0.01 | 5029 | 0.0003 | 0.0106 (0.0117) | 0.7604 |
|  |  | OCD | 0.001 | 602 | 0.0013 | −0.0219 (0.0118) | 0.2121 |
|  |  | AN/OCD | 0.05 | 18537 | 0.0003 | −0.0099 (0.0118) | 0.8044 |
| Social phobia at age 13<sup>b,c</sup> | 30 cases,<br>2396 controls | AN | -- | -- | -- | -- | -- |
|  |  | OCD | -- | -- | -- | -- | -- |
|  |  | AN/OCD | -- | -- | -- | -- | -- |
| Generalized anxiety disorder at age 13<sup>b</sup> | 100 cases,<br>2317 controls | AN | 0.001 | 1079 | 0.0039 | 0.1701 (0.1017) | 0.2682 |
|  |  | OCD | 1 | 91289 | 0.0044 | 0.1795 (0.1022) | 0.2504 |
|  |  | AN/OCD | 0.01 | 6370 | 0.0056 | 0.2078 (0.1044) | 0.1474 |
| Latent factor – physical anxiety at age 13 | 2421 | AN | 1 | 85306 | 0.0002 | −0.0101 (0.0131) | 0.8391 |
|  |  | OCD | 0.001 | 602 | 0.0004 | −0.0130 (0.0129) | 0.7252 |
|  |  | AN/OCD | 1 | 82082 | 0.0006 | −0.0159 (0.0131) | 0.5329 |
| Latent factor – worrying at age 13 | 2421 | AN | 0.2 | 37940 | 0.0008 | 0.0227 (0.0166) | 0.4388 |
|  |  | OCD | 0.001 | 602 | 0.0029 | −0.0430 (0.0161) | **0.0307** * |
|  |  | AN/OCD | 0.2 | 43327 | 0.0016 | 0.0329 (0.0165) | 0.1440 |
| Latent factor – social phobia at age 13 | 2421 | AN | 0.3 | 48846 | 0.0011 | 0.0259 (0.0155) | 0.2680 |
|  |  | OCD | 0.001 | 602 | 0.0039 | −0.0471 (0.0152) | **0.0072** * |
|  |  | AN/OCD | 0.2 | 43327 | 0.0015 | 0.0295 (0.0156) | 0.1769 |
| Generalized anxiety disorder at age 15<sup>b,c</sup> | 154 cases,<br>1899 controls | AN | 0.1 | 24166 | 0.0036 | 0.1486 (0.0849) | 0.2354 |
|  |  | OCD | 0.1 | 23463 | 0.0013 | 0.0880 (0.0840) | 0.6895 |
|  |  | AN/OCD | 0.1 | 28752 | 0.0028 | 0.1304 (0.0840) | 0.3313 |

<sup>a</sup> Principal components 1–5 were used as covariates.
<sup>b</sup> Binary phenotype.
<sup>c</sup> Due to insufficient statistical power, any binary measure with less than 50 cases is not included in the final analysis.
\* (also bolded) Statistically significant after $P$ for regression was corrected for multiple testing via 10,000 permutations (significance threshold=0.05).
Abbreviations: PGS=polygenic score; $P_T$ =best discovery $P$ threshold; SNP=single nucleotide polymorphism; $R^2$ =Nagelkerke's pseudo- $R^2$ for total variance explained in target phenotype; $\beta$ =standardized beta regression coefficient; $SE$ =standard error; AN=anorexia nervosa; OCD=obsessive-compulsive disorder; AN/OCD=anorexia nervosa/obsessive-compulsive transdiagnostic phenotype.

### OCD and anxiety symptom phenotypes and diagnoses

***Figures 1c*** and ***1d*** show the significant results for OCD and anxiety phenotypes. In the combined sample, AN PGS predicted an increased likelihood of separation anxiety at age 7 (*P*=0.0132), higher scores for the latent factors OCD dirt/germs (*P=*0.0319), worrying at age 10 (*P*=0.0163), and social phobia (*P*=0.0380) at age 13 (***Table 2***). None of the anxiety phenotypes were predicted by OCD PGS. AN/OCD PGS predicted an increased likelihood of specific phobia at age 7 (*P*=0.0181).

In girls, AN PGS predicted an increased likelihood of separation anxiety (*P*=0.0035) at age 7 (***Table 3***). OCD PGS predicted lower scores for the latent factors worrying (*P*=0.0307) and social phobia at age 13 (*P*=0.0072). AN/OCD PGS predicted an increased likelihood of separation anxiety (*P*=0.0134) and specific phobia at age 7 (*P*=0.0378).

In boys, AN PGS predicted a higher worrying score at age 10 (*P*=0.0143), whereas neither OCD nor AN/OCD PGS predicted any anxiety-related phenotypes (***Table 4***).

**Table 4.** Prediction of eating disorder, obsessive-compulsive disorder, and anxiety symptom dimensions and diagnoses using polygenic scores in boys^a^.

| Eating Disorder Symptom Dimensions and Diagnoses |  |  |  |  |  |  |  |
| --- | --- | --- | --- | --- | --- | --- | --- |
| Phenotype | Sample size | PGS | $P_T$ | N SNPs | $R^2$ | $\beta$ (SE) | Empirical $P$ |
| Body image distortion at age 10 | 2246 | AN | 0.001 | 1080 | 0.0005 | 0.0141 (0.0136) | 0.6621 |
|  |  | OCD | 0.05 | 13924 | 0.0028 | -0.0342 (0.0136) | 0.0500 |
|  |  | AN/OCD | 0.4 | 61850 | 0.0001 | -0.0053 (0.0138) | 0.9868 |
| AN at age 14<sup>b,c</sup> | 28 cases,<br>1887 controls | AN | -- | -- | -- | -- | -- |
|  |  | OCD | -- | -- | -- | -- | -- |
|  |  | AN/OCD | -- | -- | -- | -- | -- |
| Bulimia nervosa or subthreshold bulimia nervosa at age 14<sup>b,c</sup> | 19 cases,<br>1896 controls | AN | -- | -- | -- | -- | -- |
|  |  | OCD | -- | -- | -- | -- | -- |
|  |  | AN/OCD | -- | -- | -- | -- | -- |
| Binge-eating disorder or subthreshold binge-eating disorder at age 14<sup>b,c</sup> | 8 cases,<br>1907 controls | AN | -- | -- | -- | -- | -- |
|  |  | OCD | -- | -- | -- | -- | -- |
|  |  | AN/OCD | -- | -- | -- | -- | -- |
| Eating disorders not otherwise specified or purging disorder at age 14<sup>b</sup> | 174 cases,<br>1741 controls | AN | 0.001 | 1080 | 0.0075 | 0.2019 (0.0787) | **0.0400<sup>\*</sup>** |
|  |  | OCD | 0.001 | 603 | 0.0006 | -0.0595 (0.0803) | 0.8915 |
|  |  | AN/OCD | 0.001 | 1388 | 0.0031 | 0.1334 (0.0805) | 0.2667 |
| Any threshold/subthreshold eating disorder at age 14<sup>b</sup> | 229 cases,<br>1686 controls | AN | 0.01 | 5032 | 0.0058 | 0.1701 (0.0710) | 0.0614 |
|  |  | OCD | 0.001 | 603 | 0.0001 | -0.0218 (0.0710) | 0.9973 |
|  |  | AN/OCD | 0.01 | 6385 | 0.0024 | 0.1108 (0.0719) | 0.3190 |
| Fasting at age 14<sup>b,c</sup> | 44 cases,<br>1814 controls | AN | -- | -- | -- | -- | -- |
|  |  | OCD | -- | -- | -- | -- | -- |
|  |  | AN/OCD | -- | -- | -- | -- | -- |
| Purging at age 14 | 1854 | AN | 0.01 | 5032 | 0.0001 | 0.0016 (0.0034) | 0.9773 |
|  |  | OCD | 0.1 | 23487 | 0.0011 | 0.0049 (0.0034) | 0.4181 |
|  |  | AN/OCD | 0.01 | 6385 | 0.0001 | 0.0015 (0.0034) | 0.9777 |
| Binge eating at age 14 | 1878 | AN | 0.2 | 37905 | 0.0014 | 0.0137 (0.0083) | 0.2825 |
|  |  | OCD | 1 | 91248 | 0.0006 | 0.0090 (0.0083) | 0.6657 |
|  |  | AN/OCD | 0.01 | 6385 | 0.0020 | 0.0160 (0.0083) | 0.1631 |
| Fear of weight gain at age 14 | 1850 | AN | 0.01 | 5032 | 0.0016 | 0.0193 (0.0113) | 0.2553 |
|  |  | OCD | 0.001 | 603 | 0.0007 | -0.0133 (0.0114) | 0.5988 |
|  |  | AN/OCD | 0.001 | 1388 | 0.0009 | 0.0152 (0.0115) | 0.4509 |
| Compulsive exercise at age 14 | 1787 | AN | 0.001 | 1080 | 0.0046 | 0.0376 (0.0130) | **0.0141<sup>\*</sup>** |
|  |  | OCD | 0.05 | 13924 | 0.0014 | 0.0203 (0.0131) | 0.3586 |
|  |  | AN/OCD | 0.001 | 1388 | 0.0026 | 0.0287 (0.0134) | 0.0967 |
| Pressure to lose weight at age 14 | 1853 | AN | 0.001 | 1080 | 0.0034 | 0.0804 (0.0321) | **0.0432<sup>\*</sup>** |
|  |  | OCD | 0.001 | 603 | 0.0058 | -0.1069 (0.0324) | **0.0051<sup>\*</sup>** |
|  |  | AN/OCD | 0.001 | 1388 | 0.0047 | 0.0974 (0.0329) | **0.0122<sup>\*</sup>** |
| Restraint at age 14 | 1817 | AN | 0.2 | 37905 | 0.0038 | 0.0547 (0.0207) | **0.0312<sup>\*</sup>** |
|  |  | OCD | 0.05 | 13924 | 0.0010 | 0.0279 (0.0206) | 0.4822 |
|  |  | AN/OCD | 0.3 | 54002 | 0.0046 | 0.0605 (0.0208) | **0.0148<sup>\*</sup>** |
| Emotional eating at age 14 | 1760 | AN | 0.5 | 65261 | 0.0028 | 0.2485 (0.1119) | 0.0890 |
|  |  | OCD | 0.001 | 603 | 0.0017 | -0.1911 (0.1103) | 0.2695 |
|  |  | AN/OCD | 0.5 | 67970 | 0.00191 | 0.2054 (0.1119) | 0.1857 |
| External eating at age 14 | 1540 | AN | 0.001 | 1080 | 0.0006 | 0.0854 (0.0863) | 0.6987 |
|  |  | OCD | 0.01 | 3913 | 0.0014 | 0.1299 (0.0877) | 0.3993 |
|  |  | AN/OCD | 0.001 | 1388 | 0.0007 | 0.0949 (0.0887) | 0.6259 |
| Thin ideal internalization at age 14 | 1770 | AN | 0.001 | 1080 | 0.0006 | 0.0736 (0.0699) | 0.6471 |
|  |  | OCD | 0.001 | 603 | 0.0017 | −0.1216 (0.0707) | 0.2694 |
|  |  | AN/OCD | 0.01 | 6385 | 0.0011 | −0.1013 (0.0711) | 0.3837 |
| Body dissatisfaction at age 14 | 1872 | AN | 0.01 | 5032 | 0.0031 | 0.3898 (0.1616) | 0.0534 |
|  |  | OCD | 0.001 | 603 | 0.0015 | −0.273 (0.1629) | 0.2809 |
|  |  | AN/OCD | 0.3 | 54002 | 0.0022 | 0.3312 (0.1636) | 0.1324 |
| Weight and shape concern at age 14 | 1865 | AN | 0.01 | 5032 | 0.0018 | 0.0614 (0.0338) | 0.2026 |
|  |  | OCD | 0.001 | 603 | 0.0006 | −0.0366 (0.0339) | 0.6651 |
|  |  | AN/OCD | 0.5 | 67970 | 0.0009 | 0.0442 (0.0344) | 0.4775 |
| AN at age 16<sup>b,c</sup> | 15 cases,<br>1476 controls | AN | -- | -- | -- | -- | -- |
|  |  | OCD | -- | -- | -- | -- | -- |
|  |  | AN/OCD | -- | -- | -- | -- | -- |
| Bulimia nervosa at age 16<sup>b,c</sup> | 25 cases,<br>1466 controls | AN | -- | -- | -- | -- | -- |
|  |  | OCD | -- | -- | -- | -- | -- |
|  |  | AN/OCD | -- | -- | -- | -- | -- |
| Binge-eating disorder at age 16<sup>b,c</sup> | 10 cases,<br>1481 controls | AN | -- | -- | -- | -- | -- |
|  |  | OCD | -- | -- | -- | -- | -- |
|  |  | AN/OCD | -- | -- | -- | -- | -- |
| Eating disorders not otherwise specified or purging disorder at age 16<sup>b</sup> | 236 cases,<br>1255 controls | AN | 0.2 | 37905 | 0.0009 | 0.0648 (0.0723) | 0.7575 |
|  |  | OCD | 0.1 | 23487 | 0.0074 | 0.1821 (0.0721) | **0.0479\*** |
|  |  | AN/OCD | 0.001 | 1388 | 0.0008 | 0.0599 (0.0711) | 0.7776 |
| Any threshold/subthreshold eating disorder at age 16<sup>b</sup> | 286 cases,<br>1205 controls | AN | 0.2 | 37905 | 0.0008 | 0.0596 (0.0670) | 0.7633 |
|  |  | OCD | 1 | 91248 | 0.0028 | 0.1060 (0.0659) | 0.3226 |
|  |  | AN/OCD | 0.2 | 43348 | 0.0012 | 0.0708 (0.0660) | 0.6129 |
| Fasting at age 16 | 1407 | AN | 0.2 | 37905 | 0.0017 | 0.0133 (0.0085) | 0.3145 |
|  |  | OCD | 0.001 | 603 | 0.0008 | −0.0088 (0.0082) | 0.6764 |
|  |  | AN/OCD | 0.001 | 1388 | 0.0007 | −0.0085 (0.0084) | 0.6552 |
| Purging at age 16 | 1412 | AN | 0.2 | 37905 | 0.0013 | 0.0063 (0.0047) | 0.4416 |
|  |  | OCD | 0.01 | 3913 | 0.0005 | 0.0040 (0.0046) | 0.8002 |
|  |  | AN/OCD | 0.01 | 6385 | 0.0009 | 0.0052 (0.0046) | 0.5648 |
| Binge eating at age 16 | 1214 | AN | 0.05 | 15049 | 0.0010 | 0.0143 (0.0128) | 0.6055 |
|  |  | OCD | 0.2 | 38311 | 0.0023 | −0.0210 (0.0126) | 0.2894 |
|  |  | AN/OCD | 0.05 | 18541 | 0.0025 | 0.0223 (0.0127) | 0.2211 |
| Compulsive exercise at age 16 | 1346 | AN | 0.01 | 5032 | 0.0005 | 0.0154 (0.0193) | 0.8224 |
|  |  | OCD | 0.05 | 13924 | 0.0014 | 0.0268 (0.0195) | 0.4685 |
|  |  | AN/OCD | 0.001 | 1388 | 0.0008 | 0.0202 (0.0199) | 0.6562 |
| Obsessive-Compulsive Disorder Symptom Dimensions and Diagnosis |  |  |  |  |  |  |  |
| Phenotype | Sample size | PGS | P<sub>T</sub> | N SNPs | R<sup>2</sup> | β (SE) | Empirical P |
| OCD at age 10<sup>b,c</sup> | 15 cases,<br>2610 controls | AN | -- | -- | -- | -- | -- |
|  |  | OCD | -- | -- | -- | -- | -- |
|  |  | AN/OCD | -- | -- | -- | -- | -- |
| OCD latent factor – symmetry, checking at age 10 | 2607 | AN | 0.3 | 48867 | 0.0015 | 0.0207 (0.0104) | 0.1395 |
|  |  | OCD | 0.1 | 23487 | 0.0005 | −0.0123 (0.0105) | 0.6101 |
|  |  | AN/OCD | 0.1 | 28755 | 0.0013 | 0.0191 (0.0104) | 0.1879 |
| OCD latent factor – dirt/germs at age 10 | 2607 | AN | 0.001 | 1080 | 0.0023 | 0.0214 (0.0088) | 0.0503 |
|  |  | OCD | 0.01 | 3913 | 0.0002 | −0.0055 (0.0088) | 0.9378 |
|  |  | AN/OCD | 0.05 | 18541 | 0.0007 | 0.0120 (0.0088) | 0.4186 |
| OCD at age 13<sup>b,c</sup> | 6 cases,<br>2419 controls | AN | -- | -- | -- | -- | -- |
|  |  | OCD | -- | -- | -- | -- | -- |
|  |  | AN/OCD | -- | -- | -- | -- | -- |
| OCD latent factor – symmetry, checking at age 13 | 2411 | AN | 0.05 | 15049 | 0.0018 | 0.0301 (0.0145) | 0.1221 |
|  |  | OCD | 0.05 | 13924 | 0.0015 | −0.0275 (0.0144) | 0.1961 |
|  |  | AN/OCD | 0.01 | 6385 | 0.0011 | 0.0239 (0.0145) | 0.2754 |
| OCD latent factor – dirt/germs at age 13 | 2411 | AN | 0.1 | 24168 | 0.0005 | −0.0144 (0.0132) | 0.6217 |
|  |  | OCD | 0.05 | 13924 | 0.0012 | −0.0221 (0.0132) | 0.2860 |
|  |  | AN/OCD | 0.01 | 6385 | 0.0007 | −0.0175 (0.0133) | 0.4556 |
| Anxiety Symptom Dimensions and Diagnoses |  |  |  |  |  |  |  |
| Phenotype | Sample size | PGS | $P_T$ | N SNPs | $R^2$ | $\beta$ (SE) | Empirical $P$ |
| Separation anxiety at age 7<sup>b</sup> | 80 cases,<br>2670 controls | AN | 0.1 | 24168 | 0.0025 | 0.1443 (0.1141) | 0.5109 |
|  |  | OCD | 0.2 | 38311 | 0.0028 | −0.1515 (0.1138) | 0.5005 |
|  |  | AN/OCD | 0.001 | 1388 | 0.0037 | 0.1798 (0.1164) | 0.3234 |
| Specific phobia at age 7<sup>b</sup> | 51 cases,<br>2701 controls | AN | 0.01 | 5032 | 0.0121 | 0.3405 (0.1434) | 0.0567 |
|  |  | OCD | 0.1 | 23487 | 0.0025 | −0.1518 (0.1418) | 0.6784 |
|  |  | AN/OCD | 0.01 | 6385 | 0.0043 | 0.2010 (0.1427) | 0.3931 |
| Latent factor – physical anxiety at age 10 | 2607 | AN | 0.4 | 57882 | 0.0022 | 0.0341 (0.0143) | 0.0626 |
|  |  | OCD | 0.001 | 603 | 0.0005 | 0.0158 (0.0143) | 0.6611 |
|  |  | AN/OCD | 0.01 | 6385 | 0.0008 | 0.0203 (0.0145) | 0.4087 |
| Latent factor – worrying at age 10 | 2607 | AN | 0.5 | 65261 | 0.0034 | 0.0396 (0.0133) | **0.0143\*** |
|  |  | OCD | 0.5 | 68431 | 0.0006 | 0.0163 (0.0132) | 0.5582 |
|  |  | AN/OCD | 0.1 | 28755 | 0.0011 | 0.0220 (0.0133) | 0.2615 |
| Latent factor – social phobia at age 10 | 2607 | AN | 0.001 | 1080 | 0.0011 | 0.0214 (0.0124) | 0.2491 |
|  |  | OCD | 0.01 | 3913 | 0.0021 | −0.0288 (0.0124) | 0.0768 |
|  |  | AN/OCD | 0.001 | 1388 | 0.0008 | 0.0180 (0.0127) | 0.3922 |
| Social phobia at age 13<sup>b,c</sup> | 25 cases,<br>2399 controls | AN | -- | -- | -- | -- | -- |
|  |  | OCD | -- | -- | -- | -- | -- |
|  |  | AN/OCD | -- | -- | -- | -- | -- |
| Generalized anxiety disorder at age 13<sup>b</sup> | 54 cases,<br>2370 controls | AN | 0.05 | 15049 | 0.0060 | 0.2298 (0.1368) | 0.2606 |
|  |  | OCD | 0.01 | 3913 | 0.0064 | 0.2370 (0.1376) | 0.2663 |
|  |  | AN/OCD | 0.1 | 28755 | 0.0019 | 0.1277 (0.1371) | 0.7181 |
| Latent factor – physical anxiety at age 13 | 2411 | AN | 0.2 | 37905 | 0.0006 | −0.0166 (0.0141) | 0.5694 |
|  |  | OCD | 0.01 | 3913 | 0.0005 | −0.0154 (0.0139) | 0.6580 |
|  |  | AN/OCD | 0.001 | 1388 | 0.0004 | 0.0137 (0.0142) | 0.7044 |
| Latent factor – worrying at age 13 | 2411 | AN | 0.3 | 48867 | 0.0004 | 0.0159 (0.0154) | 0.6721 |
|  |  | OCD | 0.001 | 603 | 0.0003 | 0.0134 (0.0153) | 0.8070 |
|  |  | AN/OCD | 0.01 | 6385 | 0.0001 | −0.0056 (0.0154) | 0.9894 |
| Latent factor – social phobia at age 13 | 2411 | AN | 0.3 | 48867 | 0.0015 | 0.0283 (0.0148) | 0.1660 |
|  |  | OCD | 0.05 | 13924 | 0.0007 | −0.0189 (0.0146) | 0.5196 |
|  |  | AN/OCD | 0.1 | 28755 | 0.0001 | 0.0084 (0.0146) | 0.9319 |
| Generalized anxiety disorder at age 15<sup>b,c</sup> | 42 cases,<br>1813 controls | AN | -- | -- | -- | -- | -- |
|  |  | OCD | -- | -- | -- | -- | -- |
|  |  | AN/OCD | -- | -- | -- | -- | -- |

<sup>a</sup> Principal components 1–5 were used as covariates.
<sup>b</sup> Binary phenotype.
<sup>c</sup> Due to insufficient statistical power, any binary measure with less than 50 cases is not included in the final analysis.
\* (also bolded) Statistically significant after $P$ for regression was corrected for multiple testing via 10,000 permutations (significance threshold=0.05).
Abbreviations: PGS=polygenic score; $P_T$ =best discovery $P$ threshold; SNP=single nucleotide polymorphism; $R^2$ =Nagelkerke's pseudo- $R^2$ for total variance explained in target phenotype; $\beta$ =standardized beta regression coefficient; $SE$ =standard error; AN=anorexia nervosa; OCD=obsessive-compulsive disorder; AN/OCD=anorexia nervosa/obsessive-compulsive transdiagnostic phenotype.

## DISCUSSION

We were able to predict eating disorder, OCD, and anxiety phenotypes using AN, OCD, and AN/OCD PGS during different developmental points in a large population sample. In contrast to our hypothesis, the majority of symptom phenotypes were predicted by either the AN or OCD PGS and not by the AN/OCD PGS, suggesting that the genetic risk associated with these measures during development may be more disorder-specific than a transdiagnostic common factor. Alternatively, multi-trait PGS may not have reached the point at which it could enhance predictive power, especially when the contribution by one of the two disorder-specific PGS is more subtle and requires much larger sample sizes. This could also be driven by the notably smaller sample size of the OCD GWAS compared to the AN GWAS (2,688 vs 16,992 cases), therefore limiting the contribution of OCD genetic risk to the transdiagnostic PGS.

Significant PGS predictions did not fall cleanly in accordance with hypothesized disorder-specific symptom phenotypes. For instance, AN—but not OCD—PGS predicted higher scores for OCD-dirt/germs and worrying at age 10. While contamination fears are often associated with OCD, they are not unique to OCD and have a cross-disorder component. In fact, it is not uncommon for individuals with AN to present with food-related contamination fears.^54^ In our ALSPAC previous study, we found that the latent factor worrying significantly predicts eating disorder symptoms at ages 14 and 16 as well as AN diagnosis at age 16.^19^ This may suggest that uncontrolled worrying may be an underlying early symptom of AN and disordered eating that precedes the manifestation of an eating disorder. In contrast, OCD PGS predicted less worrying at age 13, which may be due to qualitative differences in worry and rumination between general anxiety and OCD.^55^ Additionally, OCD—but not AN—PGS predicted increased risk for binge eating, eating disorders not otherwise specified/purging disorder, and the presence of any eating disorder at age 16. This may be due to a larger obsessive and/or compulsive genetic component to eating disorders other than AN, which genetically has a key metabolic component.^14^ Additionally, purging often has a significant compulsive component.^56^ These results corroborate findings from our team of a contribution of OCD genetic risk to bulimia nervosa.^57^ While currently AN is the only eating disorder for which GWAS data are available, future studies on the molecular genetics of other eating disorders could shed light on their potential genetic overlap with OCD.

Our findings further emphasize the potential for sex differences in the biological pathways and vulnerabilities leading to these symptom phenotypes. As per our hypothesis, our results suggest that AN genetics may play a more prominent role in risk for eating disorder and related phenotypes in boys, as compared to girls, in *early development*. In fact, significant eating disorder symptom phenotypes at age 14 were predicted predominantly by AN PGS in boys, whereas it was the OCD PGS that predicted significant eating disorder phenotypes at age 16 in both sexes. Twin studies in female participants have shown changes in the contribution of genetic and environmental risk factors for disordered eating during different stages of adolescence—with a higher genetic risk being present during later teen years^58,59^—whereas the twin-based heritability estimate for disordered eating may be much higher in boys than girls prior to puberty (0.52 in boys versus 0 in girls),^60^ further supporting our hypothesis that AN genetic load may manifest itself earlier during adolescence in boys. Similarly, since OCD often precedes eating disorder onset temporally, it is possible that OCD genetic risk loading might need longer environmental exposure to lead to eating disorders, which could also explain why many eating disorder phenotypes were successfully predicted by OCD PGS later during development in both sexes.

Notably, AN PGS did not predict any eating disorder phenotypes in girls. It is possible that the manifestation of eating disorder phenotypes may be more heavily influenced by the modern sociocultural environment in girls than boys, therefore reducing the contribution of genetic risk. For boys, genetic risk may be of increased importance as weight-related societal pressure is less pronounced; whereas for girls, a lower genetic risk load coupled with sociocultural factors may suffice for the expression of eating disorder-related symptoms. Additionally, it is unclear how much the genetic etiology of AN overlaps with other eating disorder diagnoses or symptoms. Twin studies have shown disordered eating symptoms to be moderately heritable (i.e., a heritability estimate of up to 0.65),^59-62^ but while these symptom phenotypes have a notable twin-based genetic correlation with eating disorders other than AN, their genetic correlation with AN is much weaker (0.52 for other eating disorders versus 0.26 for AN).^62^ Although SNP-based heritability data for non-AN eating disorder phenotypes are limited and based on small sample sizes, a recent GWAS by our group did not find a significant genetic correlation between AN and bulimia factor score,^63^ despite both phenotypes showing significant SNP-based heritability estimates.^64^ While these studies were conducted in female participants only and cannot be necessarily generalized to men, currently little evidence exists at a genetic level on whether these intermediate phenotypes truly lie on a continuum with AN diagnosis.

We also observed sex differences in the genetic prediction of anxiety symptoms and diagnoses. AN PGS predicted separation anxiety at age 7 in girls and increased worrying at age 10 in boys. Epidemiological studies show over a 10-fold increase in AN risk among girls with separation anxiety disorder,^65^ and a twin-based study reported a shared genetic effect influencing liability to AN, separation anxiety, and childhood overanxious disorder (which is very similar to generalized anxiety disorder in adults) during different stages of development,^66^ supporting our findings about the presence of a shared genetic pathway between anxiety and AN. We unexpectedly observed that *lower* OCD-specific genetic risk predicted higher anxiety symptoms in girls (e.g., worrying and social phobia latent factors at age 13). As worrying and social phobia are symptoms that are shared among many anxiety disorders and may be non-specific, enhanced risk for these symptoms may arise through more general pathways. Additionally, while anxiety symptoms are common in patients with OCD, our results suggest that OCD may be distinct from anxiety disorders at a genetic level, especially for men. Replication of these associations is crucial to better understand the nature of this relationship and the importance of sex differences in the biological pathways associated with anxiety risk.

Compulsive exercise was the only intermediate phenotype that was positively predicted by more than one PGS for both sexes, strongly suggesting it may be one intermediate phenotype that, although commonly associated with eating disorders, is truly transdiagnostic, not sex-specific, and influenced by genetic risk for both AN and OCD. Together with evidence for shared genetic risk between a broad AN phenotype and general propensity for physical activity,^14^ this finding suggests that genetic factors may be particularly relevant to understanding the development of compulsive exercise in eating disorders.

Compulsive exercise encompasses many of the hallmark symptoms of AN (e.g., weight and shape concern) and OCD (e.g., compulsive behavior).^67^ Furthermore, comorbid OCD symptoms are especially pronounced in the subpopulation of AN patients with compulsive exercise,^67-71^ which has significant clinical relevance since the presence of compulsive exercise in AN is an established predictor of negative treatment outcomes, including higher pathology at discharge from inpatient treatment,^72^ relapse,^73^ and greater energy requirements for weight gain.^74^ Treatments for this symptom are currently lacking, and our results point to the need for additional investigation of the habitual and compulsive nature of exercise behavior, which may lead to targeted intervention development for this symptom that derives from a modern biobehavioral understanding of both eating disorders and OCD.

Our study has notable strengths that merit consideration. This is the first study to use AN, OCD, or AN/OCD PGS to predict eating disorder and anxiety intermediate phenotypes, and to examine sex differences in how genetic risk manifests in the presence of these intermediate phenotypes during different stages of development in a large population sample. We augmented the diagnostic approach by including intermediate phenotypes measured continuously to capture the full range of these underlying traits in the general population. Furthermore, studying these associations in the general population allows a finer understanding of intermediate phenotypes and broader psychopathology as treatment-seeking individuals might have notable differences from the general population (e.g., increased comorbid psychopathology).

Limitations of our study include reliance on self- or parent-report symptoms instead of clinical diagnoses, phenotype data being available for only a subset of participants with the potential for response bias, and potential Type 2 error due to lack of statistical power for PGS in the prediction of genetic risk. Of note, few participants met diagnostic criteria for AN, bulimia nervosa, or binge-eating disorder, whereas we had better statistical power for the non-specific eating disorder statuses, which likely explains why AN PGS did not predict AN diagnosis. Similarly, a high probability (50% or higher) for anxiety disorder diagnoses was uncommon despite our attempt to increase power through dichotomizing these items, and even with dichotomizing, we did not have enough cases to include OCD diagnosis in our outcomes. Additionally, we did not address the presence of comorbid psychiatric diagnoses, therefore we cannot account for the role of comorbidities or the genetic risk associated with these additional diagnoses. However, as comorbidity is the norm and not the exception, our results are likely to capture associations that are more likely to be present in clinical and population settings, as pure forms of eating disorders and OCD are not common. From a genetic perspective, whether all of the eating disorder-related symptom phenotypes examined as a part of our study actually fall on an etiological continuum with AN is not clear.^62^ Finally, the AN PGS was constructed using a much larger GWAS than the OCD GWAS, which may have translated to OCD PGS being underpowered and the AN/OCD transdiagnostic GWAS being more heavily skewed by AN PGS than OCD PGS.

Taken together, results of our study point to significant sex differences in the manifestation of genetic risk for eating disorder and anxiety symptoms. Genetic risk associated with AN may be a stronger predictor of eating disorder symptoms in boys, whereas OCD genetic risk may play a bigger role in symptomatology in these domains for girls, and may increase in effect across adolescence, which is in line with previous research findings that genetic effects may play a more prominent role post-puberty for eating disorder risk in girls.^75^ These findings could have important implications for earlier detection of disordered eating and anxiety-related symptoms while being mindful of sex differences. In clinical settings, men with AN often have an earlier onset, present with lower BMI and fat mass, and score higher on weight suppression than women with AN,^37-39^ raising the possibility of a stronger genetic role in the etiology of AN in men. Our genetic results provide support for this hypothesis, especially in the context of a developmental framework. Apart from sex differences, one significant observation was that compulsive exercise may be an intermediate phenotype or clinical manifestation of shared genetic risk factors for both AN and OCD in both sexes. It is possible that compulsive exercise might be a distinct AN subphenotype, and clinical research should continue to explore habitual and compulsive processes associated with this symptom. Finally, this study opens up new avenues for a clearer understanding of biology of behaviors and intermediate phenotypes in eating disorders.

## Data Availability

Researchers can apply to the Avon Longitudinal Study of Parents and Children (ALSPAC) for data access.

## ACKNOWLEDGEMENTS

ZY was responsible for genetic study design, quality control of genotype data, genetic analyses, and manuscript preparation. KS, ELG, and LCB was responsible for study design, quality control and preparation of phenotype data, statistical analysis of phenotype data, and manuscript preparation. MH was responsible for carrying out the transdiagnostic AN/OCD GWAS. MH, JJC, MM, CAM, and GB were responsible for genetic study design, oversight of genetic analyses, and manuscript preparation. CMB oversaw and contributed to development of the research question, study design, and manuscript preparation. NM and SCZ were responsible for development of the research question and provided oversight for all aspects of the study.

We are deeply grateful to all the families who took part in this study, the midwives for their help in recruiting them, and the whole ALSPAC team, which includes interviewers, computer and laboratory technicians, clerical workers, research scientists, volunteers, managers, receptionists and nurses. The UK Medical Research Council and the Wellcome Trust (Grant ref: 217065/Z/19/Z) and the University of Bristol provide core support for ALSPAC. This publication is the work of the authors and Dr. Micali will serve as guarantor for the contents of this paper.

Dr. Micali had full access to all of the data in the study and takes responsibility for the integrity of the data and the accuracy of the data analysis.

## CONSORTIA CO-AUTHORS

### Anorexia Nervosa Genetics Initiative

Jessica H. Baker, Andrew W. Bergen, Andreas Birgegärd, Joseph M. Boden, Harry Brandt, Cynthia M. Bulik, Steven Crawford, Laramie E. Duncan, Scott Gordon, Jakob Grove, Katherine A. Halmi, Anjali K. Henders, L. John Horwood, Craig Johnson, Jennifer Jordan, Anders Juréus, Allan S. Kaplan, Walter Kaye, Martin Kennedy, Katherine M. Kirk, Mikael Landén, Janne T. Larsen, Virpi M. Leppä, Paul Lichtenstein, Nicholas G. Martin, Manuel Mattheisen, James Mitchell, Grant W. Montgomery, Preben Bo Mortensen, Melissa A. Munn-Chernoff, Claes Norring, Catherine M. Olsen, Richard Parker, John F. Pearson, Nancy L. Pedersen, Liselotte Petersen, Michael Strober, Patrick F. Sullivan, Laura M. Thornton, Tracey D. Wade, Hunna J. Watson, Thomas Werge, David C. Whiteman, D. Blake Woodside, Zeynep Yilmaz

### Eating Disorders Working Group of the PGC

Hunna J. Watson, Zeynep Yilmaz, Laura M. Thornton, Christopher Hübel, Jonathan R. I. Coleman, Héléna A. Gaspar, Julien Bryois, Anke Hinney, Virpi M. Leppä, Manuel Mattheisen, Sarah E. Medland, Stephan Ripke, Shuyang Yao, Paola Giusti-Rodríguez, Ken B. Hanscombe, Kirstin L. Purves, Roger A. H. Adan, Lars Alfredsson, Tetsuya Ando, Ole A. Andreassen, Jessica H. Baker, Wade H. Berrettini, Ilka Boehm, Claudette Boni, Vesna Boraska Perica, Katharina Buehren, Roland Burghardt, Matteo Cassina, Sven Cichon, Maurizio Clementi, Roger D. Cone, Philippe Courtet, Scott Crow, James J. Crowley, Unna N. Danner, Oliver S. P. Davis, Martina de Zwaan, George Dedoussis, Daniela Degortes, Janiece E. DeSocio, Danielle M. Dick, Dimitris Dikeos, Christian Dina, Monika Dmitrzak-Weglarz, Elisa Docampo, Laramie E. Duncan, Karin Egberts, Stefan Ehrlich, Geòrgia Escaramís, Tõnu Esko, Xavier Estivill, Anne Farmer, Angela Favaro, Fernando Fernández-Aranda, Manfred M. Fichter, Krista Fischer, Manuel Föcker, Lenka Foretova, Andreas J. Forstner, Monica Forzan, Christopher S. Franklin, Steven Gallinger, Ina Giegling, Johanna Giuranna, Fragiskos Gonidakis, Philip Gorwood, Monica Gratacos Mayora, Sébastien Guillaume, Yiran Guo, Hakon Hakonarson, Konstantinos Hatzikotoulas, Joanna Hauser, Johannes Hebebrand, Sietske G. Helder, Stefan Herms, Beate Herpertz-Dahlmann, Wolfgang Herzog, Laura M. Huckins, James I. Hudson, Hartmut Imgart, Hidetoshi Inoko, Vladimir Janout, Susana Jiménez-Murcia, Antonio Julià, Gursharan Kalsi, Deborah Kaminská, Jaakko Kaprio, Leila Karhunen, Andreas Karwautz, Martien J. H. Kas, James L. Kennedy, Anna Keski-Rahkonen, Kirsty Kiezebrink, Youl-Ri Kim, Lars Klareskog, Kelly L. Klump, Gun Peggy S. Knudsen, Maria C. La Via1, Stephanie Le Hellard, Robert D. Levitan, Dong Li, Lisa Lilenfeld, Bochao Danae Lin, Jolanta Lissowska, Jurjen Luykx, Pierre J. Magistretti, Mario Maj, Katrin Mannik, Sara Marsal, Christian R. Marshall, Morten Mattingsdal, Sara McDevitt, Peter McGuffin, Andres Metspalu, Ingrid Meulenbelt, Nadia Micali, Karen Mitchell, Alessio Maria Monteleone, Palmiero Monteleone, Melissa A. Munn-Chernoff, Benedetta Nacmias, Marie Navratilova, Ioanna Ntalla, Julie K. O’Toole, Roel A. Ophoff, Leonid Padyukov, Aarno Palotie, Jacques Pantel, Hana Papezova, Dalila Pinto, Raquel Rabionet, Anu Raevuori, Nicolas Ramoz, Ted Reichborn-Kjennerud, Valdo Ricca, Samuli Ripatti, Franziska Ritschel, Marion Roberts, Alessandro Rotondo, Dan Rujescu, Filip Rybakowski, Paolo Santonastaso, André Scherag, Stephen W. Scherer, Ulrike Schmidt, Nicholas J. Schork, Alexandra Schosser, Jochen Seitz, Lenka Slachtova, P. Eline Slagboom, Margarita C. T. Slof-Op ‘t Landt, Agnieszka Slopien, Sandro Sorbi, Beata Świątkowska, Jin P. Szatkiewicz, Ioanna Tachmazidou, Elena Tenconi, Alfonso Tortorella, Federica Tozzi, Janet Treasure, Artemis Tsitsika, Marta Tyszkiewicz-Nwafor, Konstantinos Tziouvas, Annemarie A. van Elburg, Eric F. van Furth, Gudrun Wagner, Esther Walton, Elisabeth Widen, Eleftheria Zeggini, Stephanie Zerwas, Stephan Zipfel, Andrew W. Bergen, Joseph M. Boden, Harry Brandt, Steven Crawford, Katherine A. Halmi, L. John Horwood, Craig Johnson, Allan S. Kaplan, Walter H. Kaye, James E. Mitchell, Catherine M. Olsen, John F. Pearson, Nancy L. Pedersen, Michael Strober, Thomas Werge, David C. Whiteman, D. Blake Woodside, Garret D. Stuber, Scott Gordon, Jakob Grove, Anjali K. Henders, Anders Juréus, Katherine M. Kirk, Janne T. Larsen, Richard Parker, Liselotte Petersen, Jennifer Jordan, Martin Kennedy, Grant W. Montgomery, Tracey D. Wade, Andreas Birgegård, Paul Lichtenstein, Claes Norring, Mikael Landén, Nicholas G. Martin, Preben Bo Mortensen, Patrick F. Sullivan, Gerome Breen, Cynthia M. Bulik.

### Obsessive Compulsive Disorder Working Group of the PGC

Paul D. Arnold, Kathleen D. Askland, Cristina Barlassina, Laura Bellodi, O. J. Bienvenu, Donald Black, Michael Bloch, Helena Brentani, Christie L. Burton, Beatriz Camarena, Carolina Cappi, Danielle Cath, Maria Cavallini, David Conti, Edwin Cook, Vladimir Coric, Bernadette A. Cullen, Danielle Cusi, Lea K. Davis, Richard Delorme, Damiaan Denys, Eske Derks, Valsamma Eapen, Christopher Edlund, Lauren Erdman, Peter Falkai, Martijn Figee, Abigail J. Fyer, Daniel A Geller, Fernando S. Goes, Hans Grabe, Marcos A. Grados, Benjamin D. Greenberg, Edna Grünblatt, Wei Guo, Gregory L. Hanna, Sian Hemmings, Ana G. Hounie, Michael Jenicke, Clare Keenan, James Kennedy, Ekaterina A. Khramtsova, Anuar Konkashbaev, James A. Knowles, Janice Krasnow, Cristophe Lange, Nuria Lanzagorta, Marion Leboyer, Leonhard Lennertz, Bingbin Li, K-Y Liang, Christine Lochner, Fabio Macciardi, Brion Maher, Wolfgang Maier, Maurizio Marconi, Carol A. Mathews, Manuel Mattheisen, James T. McCracken, Nicole C. McLaughlin, Euripedes C. Miguel, Rainald Moessner, Dennis L. Murphy, Benjamin Neale, Gerald Nestadt, Paul Nestadt, Humberto Nicolini, Ericka Nurmi, Lisa Osiecki, David L. Pauls, John Piacentini, Danielle Posthuma, Ann E. Pulver, H-D Qin, Steven A. Rasmussen, Scott Rauch, Margaret A. Richter, Mark A. Riddle, Stephan Ripke, Stephan Ruhrmann, Aline S. Sampaio, Jack F. Samuels, Jeremiah M. Scharf, Yin Yao Shugart, Jan Smit, Daniel Stein, S. Evelyn Stewart, Maurizio Turiel, Homero Vallada, Jeremy Veenstra-VanderWeele, Michael Wagner, Susanne Walitza, Y. Wang, Jens Wendland, Nienke Vulink, Dongmei Yu, Gwyneth Zai.

